# Perineural invasion as a candidate prognostic marker beyond AJCC 8 staging in resected duodenal adenocarcinoma: a single-center retrospective cohort study

**DOI:** 10.64898/2026.07.09.26357415

**Authors:** Yong-Ping Lian, Yu-Jun Wu, Xin-Yi Chen, Xiao-Xiao Luo

## Abstract

**Background:** Duodenal adenocarcinoma (DA) is a rare gastrointestinal malignancy with limited DA-specific evidence on the prognostic role of perineural invasion (PNI) and the performance of AJCC 8th-edition staging. We described PNI, AJCC 8 discrimination, and adjuvant-chemotherapy subgroup associations in a single-center cohort.

**Methods:** We retrospectively analyzed 51 patients with curatively resected, histologically invasive DA (2013-2025). Overall survival (OS) and disease-free survival (DFS) were estimated by Kaplan-Meier analysis; prognostic associations by Cox regression. Model discrimination was quantified by the C-index; adjuvant-chemotherapy associations were explored across pre-specified subgroups. Analyses were exploratory and hypothesis-generating.

**Results:** Median follow-up was 34.9 months; 26 patients (51%) died and 32 (63%) recurred, with the highest recurrence frequency 6-12 months after surgery. AJCC 8 staging discriminated modestly (C-index 0.595); no adjacent stage pair differed significantly after Bonferroni correction (minimum raw pairwise P = 0.051). Among candidate factors, PNI had the largest effect estimate (univariable HR 2.08, 95% CI 0.88-4.90, P = 0.095) and the largest incremental discrimination when added to a stage + T + N base model (delta C-index +0.062; likelihood-ratio P = 0.145), exceeding the increments from tumor size (+0.046), differentiation (+0.034), and sex (+0.016). Adjuvant chemotherapy was associated with hazard-ratio reductions in PNI-positive (HR 0.19, 95% CI 0.02-1.59), stage III (HR 0.36, 95% CI 0.12-1.15), and node-positive (HR 0.36, 95% CI 0.12-1.15) subgroups; none reached statistical significance, consistent with limited power in subgroups of 8-22 patients.

**Conclusion:** In this small single-center cohort, PNI showed the largest prognostic effect estimate among candidate variables and the largest incremental discrimination beyond AJCC 8 stage, although neither reached statistical significance and AJCC 8 discriminated only modestly. These hypothesis-generating findings - directionally concordant with larger published DA series - support formal evaluation of PNI as a stratification variable in multicenter cohorts and in individual-patient-data meta-analyses of duodenal adenocarcinoma, rather than a change in current practice.

## Introduction

Duodenal adenocarcinoma (DA) is a rare gastrointestinal malignancy, representing approximately 0.5% of all gastrointestinal tumors yet accounting for more than 50% of small bowel adenocarcinomas [1,2,27,28]. Because of its rarity, most published series have either pooled DA with small bowel or periampullary adenocarcinomas, or relied on small single-center cohorts (n = 20-100), which has diluted DA-specific evidence regarding prognostic factors and treatment [3,4]. Surgical resection - typically pancreaticoduodenectomy for proximal (D1-D2) lesions and segmental resection for distal (D3-D4) lesions - remains the only potentially curative treatment [5]. Despite curative-intent surgery, reported 5-year overall survival has varied widely from 30% to 60%, and more than half of patients eventually experience recurrence [3,4,29,32]. The role of adjuvant chemotherapy in resected DA remains incompletely defined, and current guidelines provide limited guidance on patient selection in the absence of prospective randomized evidence [6].

Across multiple gastrointestinal and non-gastrointestinal cancers - including pancreatic ductal adenocarcinoma, distal cholangiocarcinoma, ampullary adenocarcinoma, prostate, head and neck, gastric, and breast cancers - perineural invasion (PNI), defined as the infiltration of tumor cells along nerve sheaths, has been consistently identified as an adverse prognostic factor [7,8]. In a 121-patient periampullary carcinoma series, van Roest and colleagues demonstrated that perineural growth was a more important prognostic factor than tumor localization itself [9]. Single-center data also suggest PNI has prognostic value in DA specifically - most directly the 103-patient series of Cecchini et al., in which PNI outperformed nodal involvement on multivariable analysis [10] - potentially through tumor-nerve interactions involving neurotrophic factors and axon-guidance pathways [11,12]. However, DA-specific evidence remains scarce and confined to a handful of single-center series, and whether PNI provides prognostic information beyond AJCC 8 staging in DA remains incompletely characterized.

The AJCC 8th edition staging system for small intestinal adenocarcinoma updated nodal classification to align with colon cancer (N1: 1-2 positive nodes; N2: >= 3 positive nodes) [13]. External validation in a SEER cohort of nearly 3,000 patients, however, reported only modest prognostic discrimination (C-index 0.660), with the stage IIB versus IIIA pairwise comparison failing to reach statistical significance [14]. Data on adjuvant-chemotherapy benefit in DA are likewise conflicting, plausibly in part because most studies do not stratify on adverse pathological features such as PNI [15]. In this single-center retrospective cohort of 51 patients with curatively resected DA followed for up to 12 years, we described the prognostic value of PNI, the discrimination of AJCC 8 staging, and adjuvant-chemotherapy associations across subgroups, as an exploratory, hypothesis-generating analysis intended to inform larger multicenter and meta-analytic efforts.

## Materials and methods

### Patient cohort and inclusion criteria

We retrospectively reviewed consecutive patients with histologically confirmed primary duodenal adenocarcinoma (DA) who underwent curative-intent surgical resection at our institution between January 2013 and November 2025. Inclusion criteria were: (i) age >= 18 years; (ii) pathologically confirmed **invasive** primary DA (non-ampullary); (iii) R0 or R1 resection with regional lymph node dissection; (iv) no neoadjuvant chemotherapy or radiotherapy; and (v) availability of complete clinicopathologic and follow-up data. Patients with M1 disease detected before or during surgery were not categorically excluded but were analyzed separately for staging purposes. Patients with concurrent or prior malignancies, ampullary or pancreatic primaries, R2 resection, follow-up < 3 months, or **high-grade dysplasia without histologically invasive carcinoma** were excluded. This study was approved by the Institutional Review Board of Tongji Hospital, Tongji Medical College, Huazhong University of Science and Technology (approval no. TJ-IRB20240616); given the retrospective design and use of de-identified data, written informed consent was waived. Reporting follows the STROBE statement.

### Variables and pathological assessment

Clinicopathologic data were extracted from electronic medical records and pathology reports by two reviewers independently, with discrepancies resolved by consensus. Tumor variables included location (D1 bulb; D2 descending; D3 horizontal; D4 ascending; or anastomosis), maximum diameter, differentiation (well, moderate, moderate-poor, poor), depth of invasion (T1-T4) and nodal status (N0-N2) per the 8th-edition AJCC small-intestine classification [13]. For the duodenum, invasion of the pancreas or bile duct was classified as T4 per the AJCC duodenum-specific rule. Perineural invasion (PNI) was defined as tumor cells within any layer of the nerve sheath or perineural space, per Cecchini et al. [10]. Lymphovascular invasion (LVI) was defined as identifiable tumor emboli within vascular or lymphatic spaces. Examined and metastatic lymph node counts, resection margin, and preoperative CEA (< 5 ng/mL) and CA19-9 (< 37 U/mL) were recorded.

### AJCC 8 staging and handling of unclassified N status

Stage was assigned per AJCC 8th edition. Where nodes were positive but N was otherwise unclassified, N category was inferred from the positive-node count (N1: 1-2; N2: >= 3); cases with no nodes dissected and no positive nodes were treated as N0, with a sensitivity analysis confirming this did not alter the principal findings.

### Treatment and follow-up

Procedures included pancreaticoduodenectomy, pancreas-sparing duodenectomy, or segmental duodenal resection by tumor location and depth. Adjuvant chemotherapy (commonly FOLFOX, CAPEOX, S-1, or capecitabine) and selective adjuvant radiotherapy (R1/close margins) followed contemporary practice. Follow-up was every 3-6 months for 2 years then annually, with clinical examination, tumor markers, and contrast-enhanced CT. The primary outcome was overall survival (OS, surgery to death from any cause; survivors censored at last follow-up); the secondary outcome was disease-free survival (DFS, surgery to first recurrence, death, or last follow-up). Follow-up cutoff was November 2025.

### Statistical analysis

All eligible consecutive patients were included; no a priori sample-size calculation was performed given DA rarity and the descriptive nature of this cohort. Missing data were handled by complete-case analysis (extent reported in Results, e.g., LVI in 30 of 51). Continuous variables are median (IQR); categorical as n (%). Survival was estimated by Kaplan-Meier and compared by log-rank; pairwise stage comparisons used Bonferroni correction. Univariable and multivariable Cox regression identified prognostic associations. Discrimination of nested Cox models was quantified by Harrell’s C-index; the incremental discrimination of adding PNI to a base model (stage + T + N) was assessed by the change in C-index and the likelihood-ratio test. PNI and adjuvant-chemotherapy subgroup analyses used stratified Cox regression. All tests were two-sided (alpha = 0.05). Given the sample size and multiplicity, analyses are exploratory; associations are reported with effect estimates and confidence intervals rather than emphasized dichotomous significance. Analyses used Python 3.13 (lifelines 0.30.3) and R 4.3.

## Results

### Patient characteristics and outcomes

Fifty-one patients with curatively resected invasive DA were included; one additional resected lesion showing high-grade intraepithelial neoplasia without invasion was excluded. The cohort comprised 32 men (63%) and 19 women (37%), with a median tumor size of 3.0 cm (IQR 2.0-4.0; n = 50) and the descending duodenum the most common location (33%). Disease was advanced: 88% had T3-T4 lesions, 45% had nodal involvement (N1-N2), and 73% were AJCC 8 stage IIB or higher. PNI was present in 24 patients (47%) and LVI in 8 of 21 evaluable patients (not assessed in 30). Adjuvant chemotherapy was given to 23 of 45 evaluable patients (51%) and adjuvant radiotherapy to 11 of 50 (22%). Median follow-up was 34.9 months; 26 patients (51%) died and 32 (63%) recurred (3 lost to follow-up). Median OS was 46.1 months and median DFS 14.6 months.

### Recurrence patterns

Among the 51 patients, 32 (63%) recurred: 16 (31%) local only, 14 (27%) distant only, and 2 (4%) both. Median time to recurrence was 12.5 months, with the highest recurrence frequency 6-12 months after surgery. The most common recurrence sites were retroperitoneal lymph nodes (n = 9), liver (n = 8), bone (n = 6), lung (n = 3), and anastomosis (n = 3). Post-recurrence OS varied by recurrence pattern (log-rank P = 0.003). DFS by PNI status showed no significant difference (log-rank P = 0.543).

### Survival landscape and AJCC 8 discrimination

Kaplan-Meier analysis of OS by AJCC 8 stage collapsed to three groups (I; II = IIA+IIB; III = IIIA+IIIB) showed log-rank P = 0.481. Median OS was 76.1 months for stage I (n = 6, 2 deaths), 59.7 months for stage II (n = 21, 10 deaths), and 23.7 months for stage III (n = 23, 13 deaths). DFS by stage showed log-rank P = 0.190. Pairwise OS comparisons among the five individual stages (I, IIA, IIB, IIIA, IIIB) reached significance in no pair after Bonferroni correction (minimum raw P = 0.051, I vs IIB). The C-index for AJCC 8 stage predicting OS was 0.595; for T category alone 0.577 and for N category alone 0.553.

### PNI as a prognostic factor

In the overall cohort, OS by PNI status showed log-rank P = 0.088 (PNI-negative n = 27, PNI-positive n = 24); DFS by PNI was not significantly different (P = 0.543). In the stage I-II subset, OS by PNI showed a non-significant trend (log-rank P = 0.071); in the stage III subset there was no difference (P = 0.575).

In univariable Cox regression of candidate prognostic factors, PNI had the largest effect estimate (HR 2.08, 95% CI 0.88-4.90, P = 0.095). No other candidate factor - including T3-T4, adjuvant chemotherapy, CA19-9 elevation, stage III, N+, tumor size, differentiation, sex, or CEA elevation - reached statistical significance (all P > 0.15). In pre-specified subgroup analyses, PNI effect estimates were largest in the size > 3 cm subset (HR 6.46, 95% CI 1.46-28.61, P = 0.014), the male subset (HR 3.06, 95% CI 1.03-9.12, P = 0.044), the stage I-II subset (HR 3.59, 95% CI 0.83-15.55, P = 0.087), and the N0 subset (HR 3.28, 95% CI 0.84-12.82, P = 0.088).

In nested Cox models adding each candidate to a stage + T + N base model, PNI provided the largest incremental discrimination (C-index 0.587 to 0.649; delta C-index +0.062; likelihood-ratio chi-squared = 2.13, P = 0.145), exceeding the increments from tumor size (delta C +0.046; chi-squared = 3.54, P = 0.060), differentiation (delta C +0.034; P = 0.373), and sex (delta C +0.016; P = 0.287). In a Stage + PNI hybrid grouping, median OS was 59.7 months for Early/PNI-negative (n = 13), 13.2 months for Early/PNI-positive (n = 14), and 23.7 months for stage III (n = 23); the pairwise log-rank between Early/PNI-negative and Early/PNI-positive was P = 0.071.

### Adjuvant chemotherapy associations across subgroups

In the overall cohort (n = 45 with chemotherapy data), adjuvant chemotherapy was associated with a hazard ratio of 0.60 (95% CI 0.25-1.47, P = 0.267) for OS. The largest apparent reductions were in three high-risk subsets: PNI-positive (HR 0.19, 95% CI 0.02-1.59), stage III (HR 0.36, 95% CI 0.12-1.15), and node-positive (HR 0.36, 95% CI 0.12-1.15); none reached statistical significance. Other subgroups (PNI-negative, N0, stage I-II, size strata) showed smaller or null associations; the T1-T2 subset could not be estimated (insufficient events).

## Discussion

In this single-center cohort of resected duodenal adenocarcinoma, perineural invasion (PNI) had the largest prognostic effect estimate among the candidate factors examined and the largest incremental discrimination beyond AJCC 8 stage, although neither reached statistical significance in this 51-patient series, and AJCC 8 staging itself discriminated only modestly. The consistent direction of the PNI association across all pre-specified subgroups suggests the borderline P-values reflect limited power (n = 51, 26 events) rather than absence of effect, but this remains a hypothesis to be tested in larger cohorts rather than a conclusion.

Our PNI effect estimate is directionally concordant with, though smaller and non-significant relative to, the single-center DA series of Cecchini et al. (n = 103), in which PNI was independently associated with worse OS (multivariable HR 2.52 for PNI-positive vs PNI-negative, 95% CI 1.36-4.66) and emerged as the strongest predictor [10]. Our comparable univariable effect size (HR 2.08, 95% CI 0.88-4.90) with a wider, unity-spanning interval most plausibly reflects reduced power rather than divergent biology. Notably, PNI effect estimates were numerically largest in subgroups conventionally considered lower-risk - tumors > 3 cm, stage I-II, and N0 disease - and in the stage I-II subset PNI-positive patients had a median OS (13.2 months) closer to that of stage III patients (23.7 months) than to PNI-negative early-stage patients (59.7 months). This pattern is consistent with PNI marking a biologically aggressive subset of otherwise early-stage disease, though the small subgroup sizes warrant caution.

Mechanistically, PNI reflects a reciprocal tumor-nerve crosstalk mediated by neurotrophic factors and axon-guidance molecules [11], with cross-cancer studies - none characterized specifically in DA - implicating NGF/TrkA, GDNF/RET, artemin, and related pathways [12,18,19,20]. By analogy with these data, the more aggressive behavior of PNI-positive DA than its anatomic stage alone would predict is at least biologically plausible.

In our cohort, AJCC 8 staging showed modest discrimination (C-index 0.595), closely paralleling the SEER validation of Oweira et al. (C-index 0.660; n = 2,997), where the stage IIB-versus-IIIA comparison likewise failed to reach significance [14]. That a small single-center series and a large population-based dataset converge on modest discrimination suggests a genuine ceiling for anatomic staging in this disease rather than a sample-size artefact. Adding PNI to a stage + T + N base model produced the largest incremental discrimination among the variables tested (delta C-index +0.062), though the likelihood-ratio test did not reach significance in this small sample (P = 0.145) and the increment from continuous tumor size was of similar magnitude (delta C +0.046). These observations, taken with prior DA-specific data, support the routine documentation of PNI in pathology reports and its formal evaluation as a candidate staging covariate in larger cohorts, rather than any immediate change to staging.

The role of adjuvant chemotherapy in resected DA remains incompletely defined, with conflicting data plausibly reflecting inconsistent stratification on adverse pathology [15]. In our exploratory subgroup analysis, apparent hazard-ratio reductions with adjuvant chemotherapy were concentrated in PNI-positive, stage III, and node-positive subsets, with little apparent effect in PNI-negative or T1-T2 patients; none reached significance. The direction is broadly consistent with the multicenter analysis of Zaanan et al. (n = 354), in which adjuvant-chemotherapy benefit concentrated in high-risk (T4, N2) disease, and with SEER propensity-matched data [15,16] - although that same multicenter work found no benefit in the duodenal subsite specifically, underscoring that our non-significant subgroup signals are hypothesis-generating only. A definitive answer will come from randomized data such as J-BALLAD (JCOG1502C) [17]; PNI would be a reasonable pre-specified stratification variable for biomarker-driven analyses of such trials.

This study has several limitations. First, it is a single-center retrospective cohort of 51 patients with 26 deaths; the events-per-variable constraint limits multivariable modeling, and all principal associations are statistically non-significant and exploratory. Second, LVI was assessable in only 21 of 51 cases, precluding adjustment for this correlated variable, so the signal attributed to PNI may partly reflect uncaptured LVI. Third, adjuvant-chemotherapy regimen and completion detail were incomplete in 6 of 45 evaluable patients, and molecular markers (MMR/MSI, KRAS) were available only in a subset. Fourth, the cohort spans 12 years, over which staging and treatment practices evolved. These findings should therefore be read as hypothesis-generating and, where possible, contributed as individual-patient data to multicenter analyses and meta-analyses of PNI in duodenal adenocarcinoma.

In summary, in resected duodenal adenocarcinoma, PNI showed the largest prognostic effect estimate and the largest incremental discrimination beyond AJCC 8 stage among the factors examined, without reaching statistical significance in this small cohort. Concordant with larger published series, these data support documenting PNI in DA pathology reports and evaluating it formally as a stratification variable in larger cohorts, meta-analyses, and biomarker-driven trials.

## Data Availability

All data produced in the present study are available upon reasonable request to the authors

## Figure legends

**Figure 1.**
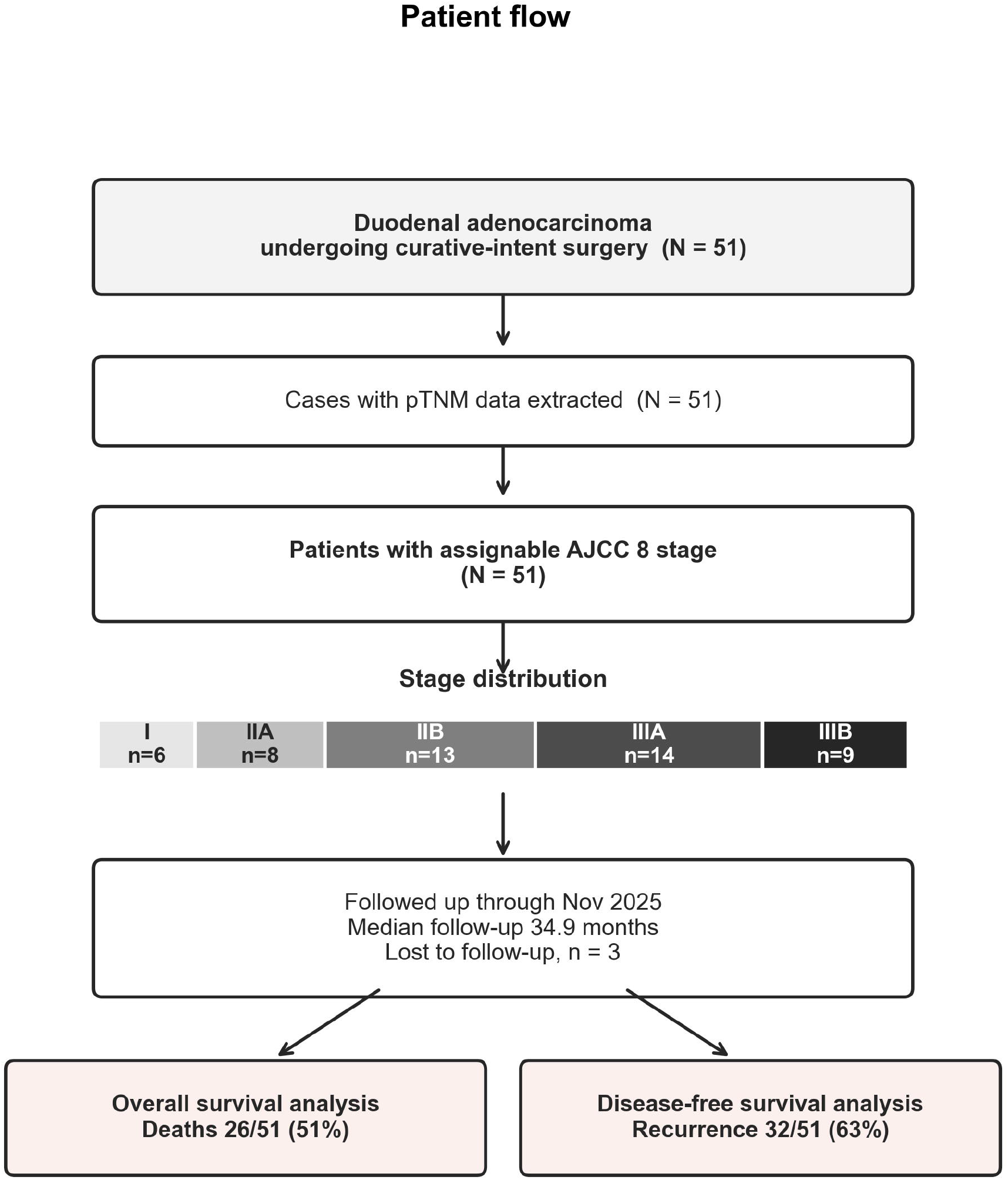
Patient flow. Selection of the 51 curatively resected invasive duodenal-adenocarcinoma patients, AJCC 8 stage distribution, follow-up, and outcome endpoints (deaths 26/51; recurrences 32/51).

**Figure 2.**
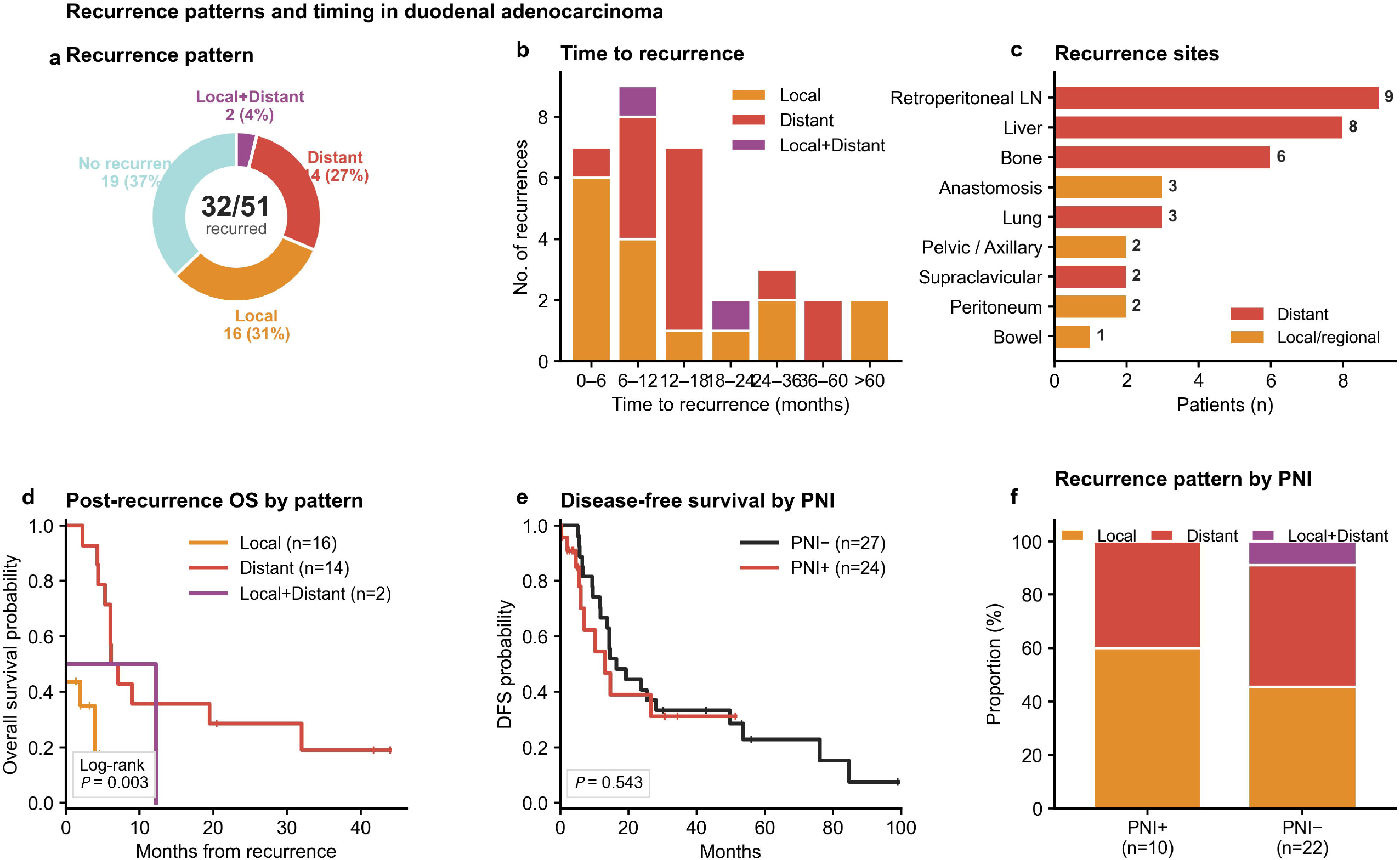
Recurrence patterns. Recurrence type (local/distant/both), timing after surgery, anatomical sites, and post-recurrence survival by pattern.

**Figure 3.**
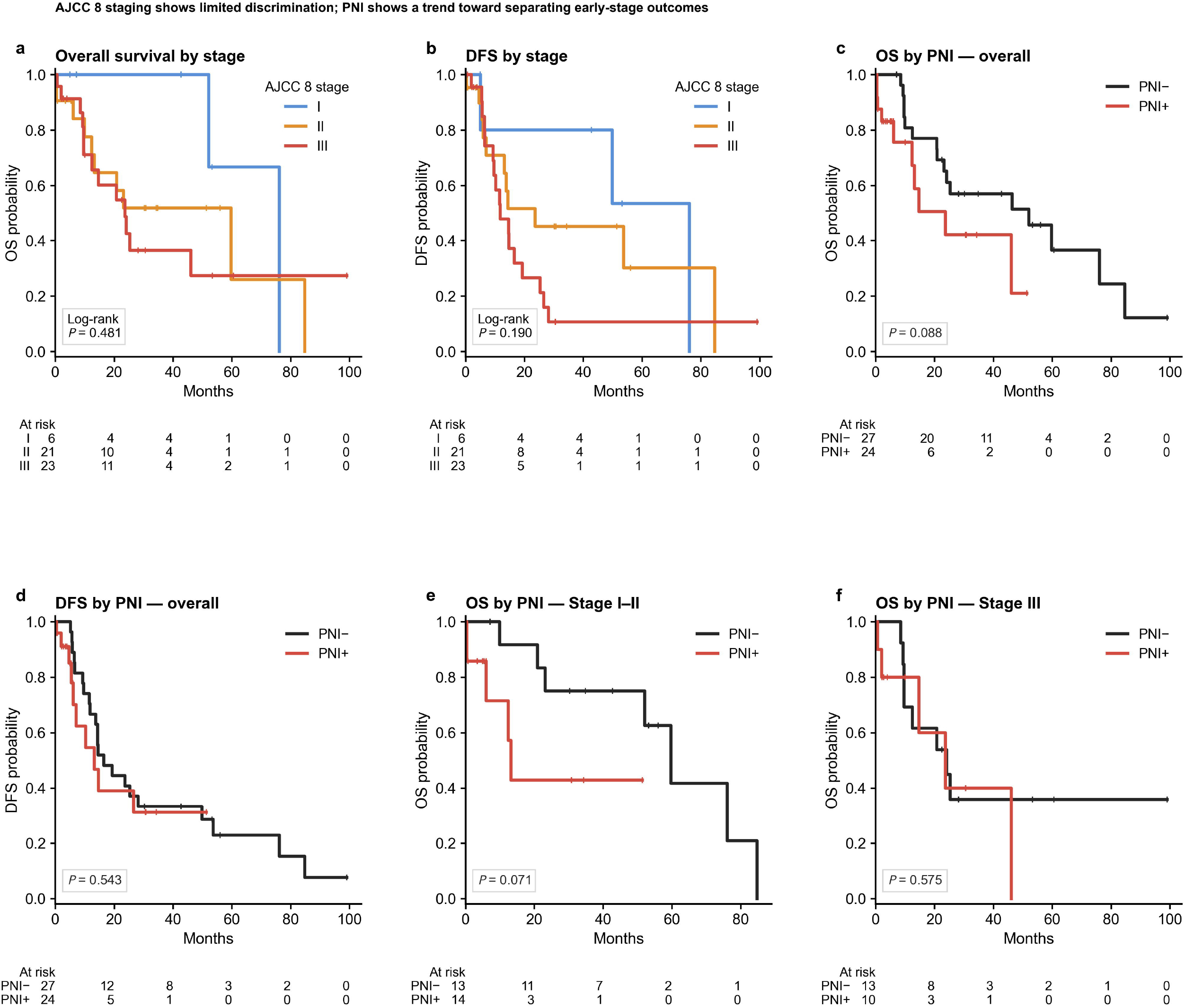
Survival landscape and AJCC 8 discrimination. Kaplan-Meier overall survival (OS) and disease-free survival (DFS) by AJCC 8 stage (three groups) and by PNI status, overall and within the stage I-II and stage III subsets, with number-at-risk tables and log-rank P-values.

**Figure 4.**
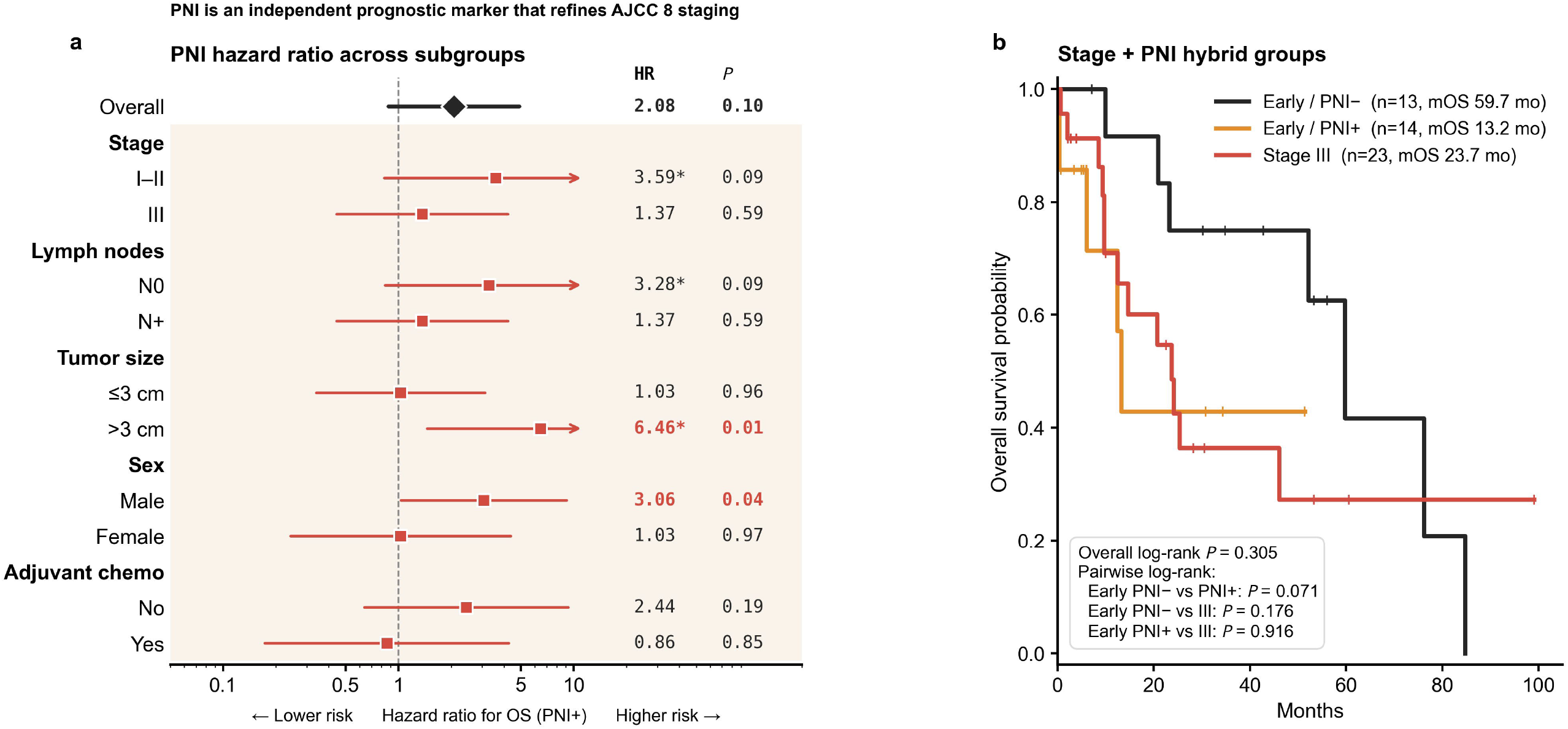
PNI as a prognostic factor. Univariable Cox forest plot of candidate prognostic factors for OS, and Kaplan-Meier OS for the Stage + PNI hybrid grouping (Early/PNI-, Early/PNI+, Stage III).

**Figure 5.**
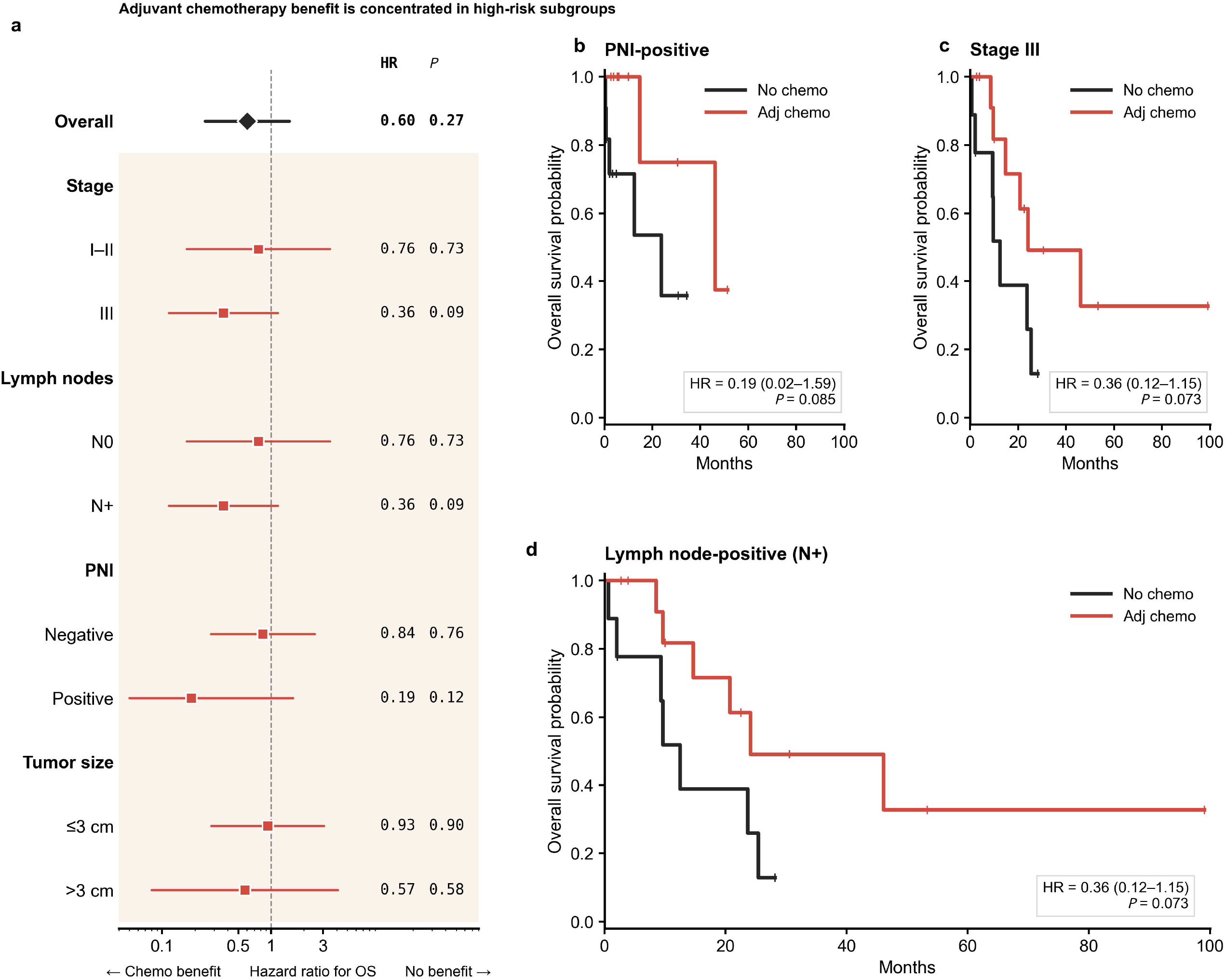
Adjuvant chemotherapy associations by subgroup. Forest plot of the adjuvant-chemotherapy hazard ratio for OS across pre-specified subgroups (overall, by stage, PNI, nodal status, T category, size, sex). All estimates are exploratory and non-significant.

## Declarations

### Ethics approval and consent to participate

Approved by the Institutional Review Board of Tongji Hospital, Tongji Medical College, Huazhong University of Science and Technology (approval no. TJ-IRB20240616). Given the retrospective design and use of de-identified data, the requirement for written informed consent was waived.

### Availability of data and materials

The de-identified individual-patient dataset supporting this study is available from the corresponding author on reasonable request and is provided as the companion individual-patient cohort for the associated aggregate meta-analysis of PNI in duodenal/small-bowel adenocarcinoma.

### Authors’ contributions

All authors contributed to study conception, data acquisition and interpretation, and critical revision of the manuscript, and approved the final version. Y-P.L. and Y-J.W. contributed equally as co-first authors; X-Y.C. and X-X.L. are co-corresponding authors.

### Declaration of generative AI in the writing process

During the preparation of this work, the authors used a large language model to assist with drafting and language editing. All content was subsequently reviewed, edited, and verified by the authors, who take full responsibility for the accuracy and integrity of the work.

## Acknowledgements

None.

## Preprint

This manuscript is posted as a preprint and has not been peer reviewed.

## References

1. Aparicio T, et al. Small Bowel Adenocarcinoma. Gastroenterol Clin North Am. 2016;45(3):447–57. PMID 27546842.

2. Bilimoria KY, et al. Small bowel cancer in the United States. Ann Surg. 2009;249(1):63–71. PMID 19106677.

3. Cloyd JM, et al. Does the extent of resection impact survival for duodenal adenocarcinoma? Ann Surg Oncol. 2015;22(2):573–80. PMID 25160736.

4. Meijer LL, et al. Outcomes and Treatment Options for Duodenal Adenocarcinoma: A Systematic Review and Meta-Analysis. Ann Surg Oncol. 2018;25(9):2681–92. PMID 29946997.

5. Solaini L, et al. Outcome after surgical resection for duodenal adenocarcinoma in the UK. Br J Surg. 2015;102(6):676–81. PMID 25776995.

6. Benson AB, et al. Small Bowel Adenocarcinoma, NCCN Guidelines v1.2020. J Natl Compr Canc Netw. 2019;17(9):1109–33. PMID 31487687.

7. Liebig C, et al. Perineural invasion in cancer: a review. Cancer. 2009;115(15):3379–91. PMID 19484787.

8. Garajova I, Giovannetti E. Targeting Perineural Invasion in Pancreatic Cancer. Cancers. 2024;16(24):4260. PMID 39766161.

9. van Roest MH, et al. Results of pancreaticoduodenectomy in periampullary adenocarcinoma: perineural growth more important than tumor localization. Ann Surg. 2008;248(1):97–103. PMID 18580212.

10. Cecchini S, et al. Superior prognostic importance of perineural invasion vs. lymph node involvement after curative resection of duodenal adenocarcinoma. J Gastrointest Surg. 2012;16(1):113–20. PMID 22005894.

11. Selvaggi F, et al. Perineural Invasion in Pancreatic Ductal Adenocarcinoma: From Molecules towards Drugs. Cancers. 2022;14(23):5793. PMID 36497277.

12. Selvaggi F, et al. Perineural invasion in pancreatic cancer: biological function in R status, prognosis, and pain. Surg Open Sci. 2025;24:58–60. PMID 40114679.

13. Amin MB, et al. The Eighth Edition AJCC Cancer Staging Manual. CA Cancer J Clin. 2017;67(2):93–99. PMID 28094848.

14. Oweira H, et al. Assessment of the external validity of the AJCC 8th staging system for small intestinal adenocarcinoma. J Gastrointest Oncol. 2019;10(3):421–28. PMID 31183191.

15. Zaanan A, et al. Adjuvant chemotherapy benefit according to T and N stage in small bowel adenocarcinoma. JNCI Cancer Spectr. 2023;7(5):pkad064. PMID 37774004.

16. Luo Y, et al. Survival benefit of adjuvant chemotherapy for duodenal adenocarcinoma: PSM analysis. Transl Cancer Res. 2025;14(12):8725–36. PMID 41510088.

17. Kitahara H, et al. Randomized phase III trial of post-operative chemotherapy for stage I/II/III small bowel adenocarcinoma (JCOG1502C, J-BALLAD). Jpn J Clin Oncol. 2019;49(3):287–90. PMID 30590606.

18. Ma J, et al. NGF and TrkA and correlation with perineural invasion in pancreatic cancer. J Gastroenterol Hepatol. 2008;23(12):1852–59. PMID 19120874.

19. Cavel O, et al. Endoneurial macrophages induce perineural invasion via GDNF/RET. Cancer Res. 2012;72(22):5733–43. PMID 22971345.

20. Ceyhan GO, et al. The neurotrophic factor artemin promotes pancreatic cancer invasion. Ann Surg. 2006;244(2):274–81. PMID 16858191.

