## Supplementary figures and images for "Perineural invasion as a candidate prognostic marker beyond AJCC 8 staging in resected duodenal adenocarcinoma: a single-center retrospective cohort study"

### supplementary FigureS1

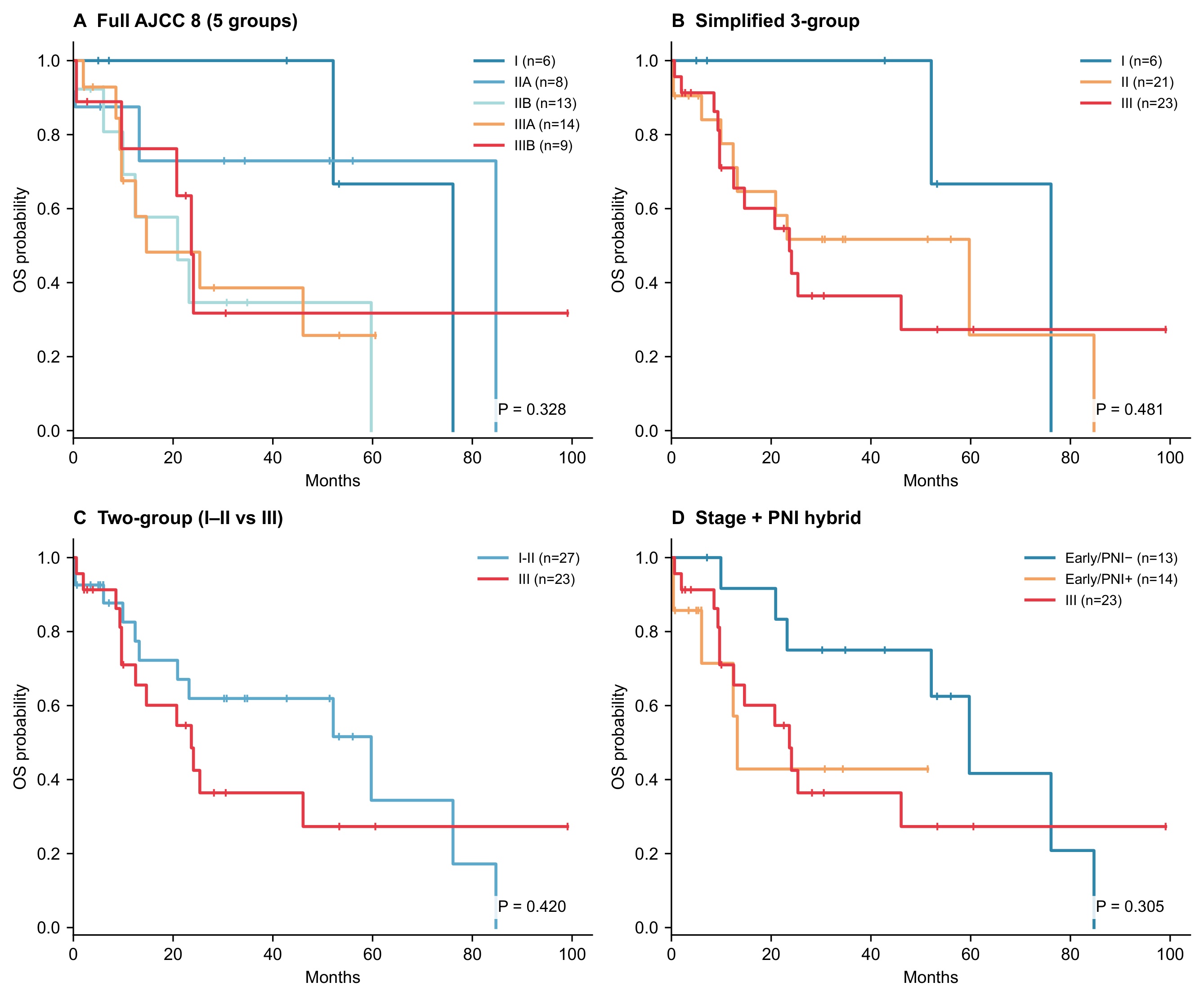
